# Anti-CD320 Autoantibodies and Central Nervous System Vitamin B12 Deficiency in Idiopathic Myelopathy

**DOI:** 10.64898/2026.01.29.26345179

**Authors:** John V. Pluvinage, David Acero-Garces, Giacomo Greco, Carson E. Moseley, Sukhman Sidhu, Kelsey C. Zorn, Sravani Kondapavulur, Megan Richie, Vanja Douglas, Sonam Mohan, John Neely, Stefano Masciocchi, Pietro Businaro, Alexis García Sarreón, Ariadna Gifreu, Krista McCutcheon, Colette Caspar, Colin Zamecnik, Asritha Tubati, Andoni I. Asencor, Madina Tugizova, Martineau Louine, Leah Zuroff, Josiah Gerdts, Mary Karalius, Alyssa Nylander, Max Liu, Iyas Daghlas, Leena Suleiman, Todd Nguyen, Benjamin Meyer, Karen Ibarra, Felicia Chow, Alexandra Galati, Yair Mina, Camilo Toro, Min Kang, Maulik Shah, Elan L. Guterman, Catherine G. Suen, Chu-Yueh Guo, Carolyn Bevan, Sarah F. Wesley, Kathryn Kvam, Sydney Lee, Ahmed Abdelhak, Thomas Martin, Yun-Han Huang, Sarah B. Berman, Jenny Linnoila, John Engstrom, Andrew McCaddon, Ari J. Green, Ralph Green, Bruce Cree, Stephen Hauser, Joseph L. DeRisi, Samuel J. Pleasure, Jeffrey M. Gelfand, Gary Álvarez Bravo, Matteo Gastaldi, Carlos A. Pardo, Michael R. Wilson

**Affiliations:** Department of Neurology, University of California, San Francisco (UCSF), San Francisco, CA, USA; Weill Institute for Neurosciences, UCSF, San Francisco, CA, USA; Department of Neurology, Johns Hopkins University School of Medicine, MD, Baltimore, MD, USA; Neuroimmunology Research Unit, IRCCS Mondino Foundation, Pavia, Italy; Department of Brain and Behavioural Sciences, University of Pavia, Pavia, Italy; Department of Biochemistry and Biophysics, UCSF, San Francisco, CA, USA; Department of Neurosurgery, UCSF, San Francisco, CA, USA; Kaiser Neurology, San Jose, CA, USA; Girona Biomedical Research Institute (IDIBGI), Neurodegeneration and neuroinflammation group, Salt, Spain; Dr. Josep Trueta University Hospital and Santa Caterina Hospital, Girona Neuroimmunology and Multiple Sclerosis Unit, Neurology Department, Girona-Salt, Spain; Neurology Department, Milford Regional Medical Center, Milford, MA, USA; Neuroscience Institute, The Queen’s Medical Center, University of Hawaii, Honolulu, Hawaii, USA; National Institute of Neurological Disorders and Stroke, National Institutes of Health (NIH), Bethesda, MD, USA; Office of the Clinical Director and Medical Genetics Branch, National Human Genome Research Institute, NIH, Bethesda, MD, USA; San Francisco VA Medical Center, San Francisco, CA, USA; Department of Neurology, Division of Neuroimmunology, Columbia University Vagelos College of Physicians and Surgeons, New York, NY, USA; Department of Neurology & Neurological, Stanford University, Stanford, CA, USA; Department of Neurology, University of Utah, Salt Lake City, UT, USA; Helen Diller Family Comprehensive Cancer Center, UCSF, San Francisco, CA, USA; Division of Rheumatology, Department of Medicine, UCSF, San Francisco, CA, USA; Department of Neurology, University of Pittsburgh, Pittsburgh, PA, USA; Faculty of Social and Life Sciences, Wrexham University, Wrexham, UK; Department of Pathology and Laboratory Medicine, University of California, Davis, CA, USA; University of Girona, Medical Sciences Department, Girona, Spain; Department of Pathology, Johns Hopkins University School of Medicine, MD, USA; Arc Institute, Palo Alto, CA, USA

## Abstract

**Background:** Disorders affecting the spinal cord (myelopathies) can cause severe disability. Despite diagnostic advances, approximately 12-18% of myelopathy cases continue to elude an etiological diagnosis, hampering effective treatment.

**Methods:** This retrospective, multicenter, tertiary care cohort study conducted from 2014 to 2025 evaluated archived biofluids from patients with IM, known autoimmune myelitis, or other neurological diseases (ONDs). Proteome-wide phage display was used to discover novel autoantibodies. Targeted immunoassays were used to screen for a candidate autoantibody. Downstream metabolites were measured in the cerebrospinal fluid (CSF).

**Results:** Autoantibodies targeting the transcobalamin receptor (CD320) responsible for cellular transport of vitamin B12 were identified in 18 out of 32 IM patients (56%) in a discovery cohort. Bioactive B12 concentration was decreased in the CSF of anti-CD320 positive patients compared to OND controls (*P* = 0.0273), indicative of autoimmune B12 central deficiency (ABCD). Compared to anti-CD320 negative IM cases, anti-CD320 positive IM cases demonstrated a higher frequency of subacute time course (56% vs 7%, *P* = 0.008), normal CSF profile (83% vs 50%, *P* = 0.044), and dorsolateral spinal cord abnormalities on magnetic resonance imaging (MRI) (61% vs 7%, *P* = 0.003). In two independent validation cohorts comprising 94 and 25 patients with IM, anti-CD320 was detected in 43 (46%) and 12 (48%) patients, respectively. Comorbid anti-CD320 was detected in a smaller proportion of patients with other known autoimmune etiologies of myelopathy. Five anti-CD320 positive IM patients received B12 supplementation with or without concurrent immunosuppression, and four out of five clinically improved.

**Conclusions:** ABCD is associated with a substantial proportion of IM. Screening for anti-CD320 followed by metabolic confirmation of a CNS-restricted B12 deficiency may be considered in the diagnostic evaluation of myelopathy.

## Introduction

Patients with myelopathy present with weakness, sensory deficits, and/or autonomic dysfunction. Etiologies can broadly be divided into non-inflammatory myelopathy (e.g. compressive, nutritional, vascular, neoplastic) and inflammatory myelitis/myelopathy (e.g. multiple sclerosis (MS), neuromyelitis optica spectrum disorder (NMOSD), myelin oligodendrocyte glycoprotein antibody-associated disease (MOGAD), sarcoidosis, rheumatologic, infectious, para-infectious, paraneoplastic).^1^ The discovery of pathogenic autoantibodies (e.g. anti-AQP4 in NMOSD) has improved the etiological diagnosis of myelopathy and led to paradigm-shifting treatments.^2,3^ However, approximately 12-18% of myelopathy cases continue to elude an etiological diagnosis despite exhaustive clinical investigation.^4,5^ Furthermore, the mechanisms of many “known” causes of myelopathy (e.g. rheumatologic, para-infectious) remain unknown.^6^ Exploring these unexplained myelopathies may reveal new etiologies amenable to targeted therapies for patients who currently endure diagnostic uncertainty and untreated disability. Here, we used proteome-wide phage display technology to explore the hypothesis that novel pathogenic autoantibodies underlie some cases of IM.

## Methods

### Study design and participants

We evaluated multiple groups of patients in an initial discovery cohort, an extended discovery cohort, and two validation cohorts (Figure 1). The initial discovery cohort comprised patients enrolled in a research study to detect novel autoantibodies in suspected neuroinflammatory disease (NID, University of California, San Francisco; IRB# 13-12236). One patient (Case 13) was previously reported.^7^ The extended discovery cohort added patients enrolled in the NID study after the initial screen as well as additional patients enrolled in a research study of myelopathy at the Unit of Neuroimmunology and Multiple Sclerosis of Girona (UNIEMTG, Comitè D’Ètica D’Investigació Amb Medicaments Girona, #1090/2015). For all patients in the initial and extended discovery cohorts, electronic medical records were reviewed to ensure completeness of information, availability of MRI images or reports, and a final diagnosis by a neurologist. Exclusion criteria for all patients in the discovery cohort were: 1) age under 18 years, 2) a final non-myelopathy diagnosis, 3) incomplete clinical information precluding diagnostic validation, or 4) a final diagnosis of a known cause of myelopathy (e.g. systemic B12 deficiency, NMOSD). Regarding exclusion criterion (4), we followed a previously published methodology^8^ to classify myelopathies by etiology with the following revisions: 1) seronegative NMOSD cases with negative anti-AQP4 and anti-MOG serologies were classified as idiopathic, 2) rheumatologic myelopathies associated with systemic lupus erythematosus (SLE) or Sjögren’s syndrome with negative anti-AQP4 and anti-MOG serologies were classified as idiopathic, and 3) para/post-infectious myelopathies without an identified causative organism were classified as idiopathic. As a control group, CSF was collected from a cohort of patients with first presentation of multiple sclerosis (MS) (no steroids within 1 month of collection, no prior disease-modifying therapy) or ONDs (migraine, NMOSD) (IRB# 14-15278, UCSF ORIGINS). The validation cohorts were retrospectively assembled from subjects enrolled at the Johns Hopkins Myelitis and Myelopathy Center (validation cohort 1, IRB# NA-00003551) or Fondazione Istituto Neurologico Nazionale C. Mondino (validation cohort 2, PV-220333) using the same inclusion and exclusion criteria as the discovery cohort. Confirmed cases of anti-AQP4+ myelitis and anti-MOG+ myelitis were included as additional controls in validation cohort 2. Detailed information on treatment regimen and clinical response was collected in the discovery and extended discovery cohorts. Time course was defined as acute (<1 month), subacute (1 month – 6 months), or chronic (>6 months).

**Figure 1.**
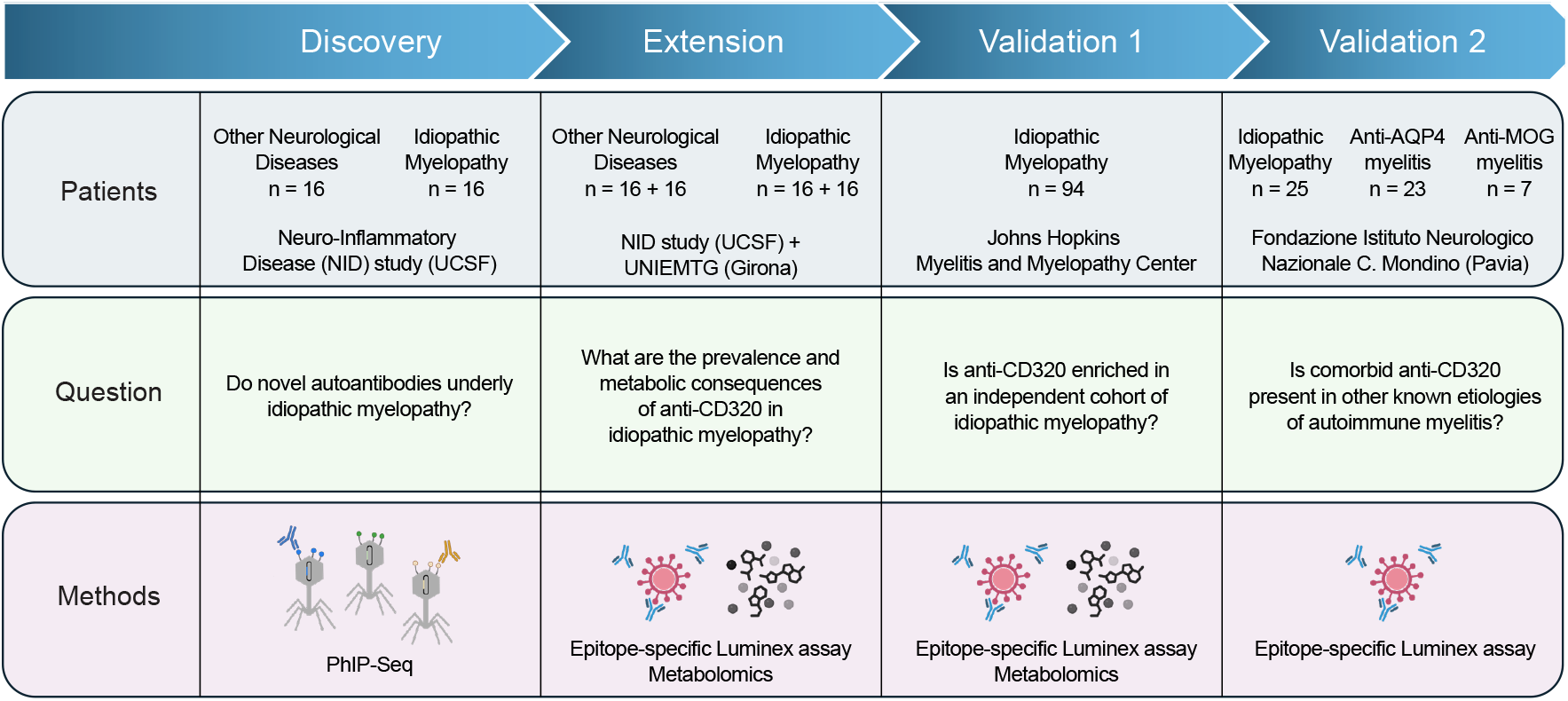
Study design and cohort summary. Multiple groups of patients were evaluated in an initial discovery cohort, an extended discovery cohort, and two validation cohorts. Patient origin, associated research question, and experimental methods used for each cohort are detailed above. For IM cases, the same inclusion and exclusion criteria were used across all cohorts (see Methods).

### Luminex assay

We adapted a previously published protocol for autoantibody detection using the Luminex platform.^9^ Briefly, a peptide comprising the epitope for anti-CD320 identified from patients with IM was synthesized and conjugated to BSA via carbodiimide coupling to a C-terminal cysteine.

Spectrally-distinct Luminex magnetic beads were conjugated to CD320 peptide-BSA or BSA alone via NHS coupling to free amines following the manufacturer’s protocol (Luminex, 40-50016). CSF was incubated with Luminex beads at a final dilution of 1:40 (CSF) or 1:500 (serum) in PBS + 0.05% Tween 20 (PBST) containing 2% non-fat milk for 1 hour at room temperature with agitation. Samples were washed two times with PBST, then stained with PE-conjugated anti-human IgG Fc secondary antibody (1:2,000, BioLegend, 637310) for 30 minutes at room temperature with agitation. Beads were washed three times with PBST and analyzed on a Luminex LX200 cytometer. Quantitative accuracy of this assay was confirmed using a standard curve with a recombinant patient-derived monoclonal anti-CD320 autoantibody.^7^ Positive samples were identified by calculating the fold change (FC) of the mean fluorescence intensities (MFI) for the CD320 peptide compared to the BSA control peptide. FC values corresponding to a concentration previously shown to functionally block B12 uptake *in vitro* were considered positive.^7^

### Cell-based assay

HEK293T cells were transfected with a FLAG-tagged CD320 construct packaged within the pCMV6 entry plasmid (Origene, RC200073) using Lipofectamine 3000 according to the manufacturer’s protocol. After 24 hours, cells were fixed with 4% PFA for 10 minutes at room temperature and stained with recombinant patient-derived anti-CD320 and rabbit anti-FLAG (CST, 14793, 1:800) at 4 degrees overnight. Secondary staining was conducted with AlexaFluor-conjugated antibodies at room temperature for 1 hour. Fluorescent images were acquired on a Zeiss LSM780 confocal laser scanning microscope.

### Metabolomics

Holotranscobalamin (bioactive B12) concentration in CSF was measured by a commercial enzyme-linked immunosorbent assay (ELISA) (Tecan, 30221798) according to the manufacturer’s protocol. Methylmalonic acid (MMA) concentration in CSF was measured by targeted gas chromatography-tandem mass spectrometry (GC-MS/MS) at Bevital AS (Norway) according to a previously published protocol.^10^

## Results

A total of 2,587 patients were enrolled in a suspected neuroinflammatory disease research study from 2014 to 2025 (Supplementary Figure 1A). This study is enriched for patients with neurologic deficits of unknown etiology despite exhaustive clinical investigation and a high suspicion for occult infectious or autoimmune causes. 200 patients (7.7%) were diagnosed with myelopathy or myelitis and confirmed upon review of the electronic medical record. 184 patients (92%) eventually received an etiological diagnosis, leaving 16 patients (8%) with the diagnosis of IM. For comparison, 16 patients were randomly selected from a cohort of OND controls comprising mainly MS (n = 14; migraine, n = 1; NMOSD, n = 1). As opposed to healthy controls, these OND controls had previously undergone lumbar puncture with bio-banked CSF readily available and demonstrated variable degrees of intrathecal inflammation.

To identify potential autoantibodies underlying IM, we performed phage immunoprecipitation sequencing (PhIP-Seq) on CSF from OND controls and IM cases (Figure 2A). We identified 20 autoantibodies enriched in the CSF of IM cases compared to OND controls (Figure 2B). Most of these autoantibodies targeted antigens with intracellular localization or low expression in the central nervous system (CNS) according to the Human Protein Atlas (proteinatlas.org),^11^ suggesting a lower likelihood of pathogenicity (Supplementary Table 1).^12,13^ However, one autoantibody enriched in IM targeted the transcobalamin receptor (CD320), a cell-surface vitamin B12 transporter highly expressed in the CNS at the blood-brain barrier.^14,15^ CSF autoantibodies from every positive case enriched the same epitope in the extracellular domain of CD320 (Supplementary Figure 2A). Antibodies targeting this epitope have previously been shown to impair B12 transport and are associated with a CNS-restricted B12 deficiency called autoimmune B12 central deficiency (ABCD).^7^

**Table 1.** Comparison of anti-CD320 negative and positive IM cases. Age, sex, MRI pattern, serum findings, CSF findings, immune-related comorbidities, and time course were compared between anti-CD320 negative and positive IM cases using a two-sided Fisher’s exact test. Age, serum B12, and serum methylmalonic acid (MMA) concentration were compared using a two-sided student’s t-test. MRI patterns classified as “other” included isolated dorsal, isolated lateral, anterior, or holocord T2 abnormalities. CSF was classified as inflammatory if there was a pleocytosis (WBC >= 6), an elevated IgG index (>0.7), or CSF-specific oligoclonal bands (>1) that were absent in the serum. CSF was not available from one patient. Autoimmune conditions included psoriatic arthritis, inflammatory bowel disease, graft-versus-host disease, post-transplant lymphoproliferative disorder, and myasthenia gravis. Time course is defined as acute (<1 month), subacute (1 month – 6 months), or chronic (>6 months).

|  | Anti-CD320 negative | Anti-CD320 positive | *P*-value |
| --- | --- | --- | --- |
| **No. of patients (%)** | 14 (44) | 18 (56) |  |
| **Age (range)** | 50 (32-71) | 57 (26-80) | 0.141 |
| **Sex (%)** |  |  | 0.473 |
| Female | 7 (50) | 6 (33) |  |
| Male | 7 (50) | 12 (67) |  |
| **MRI pattern (%)** |  |  | *0.003 |
| Normal | 3 (21) | 1 (6) |  |
| Dorsolateral | 1 (7) | 11 (61) |  |
| Other | 10 (72) | 6 (33) |  |
| **Serum findings** |  |  |  |
| B12 (pg/mL) | 745 | 655 | 0.574 |
| MMA (μM) | 0.12 | 0.18 | 0.154 |
| **CSF findings (%)** |  |  | *0.044 |
| Normal | 7 (50) | 15 (83) |  |
| Inflammatory | 7 (50) | 2 (11) |  |
| **Comorbidities (%)** |  |  | 0.999 |
| Antecedent viral illness | 3 (21) | 5 (28) |  |
| Autoimmune condition | 2 (14) | 4 (22) |  |
| HIV | 2 (14) | 2 (11) |  |
| **Time course (%)** |  |  | *0.008 |
| Acute | 2 (14) | 2 (11) |  |
| Subacute | 1 (7) | 10 (56) |  |
| Chronic | 11 (79) | 6 (33) |  |

**Figure 2.**
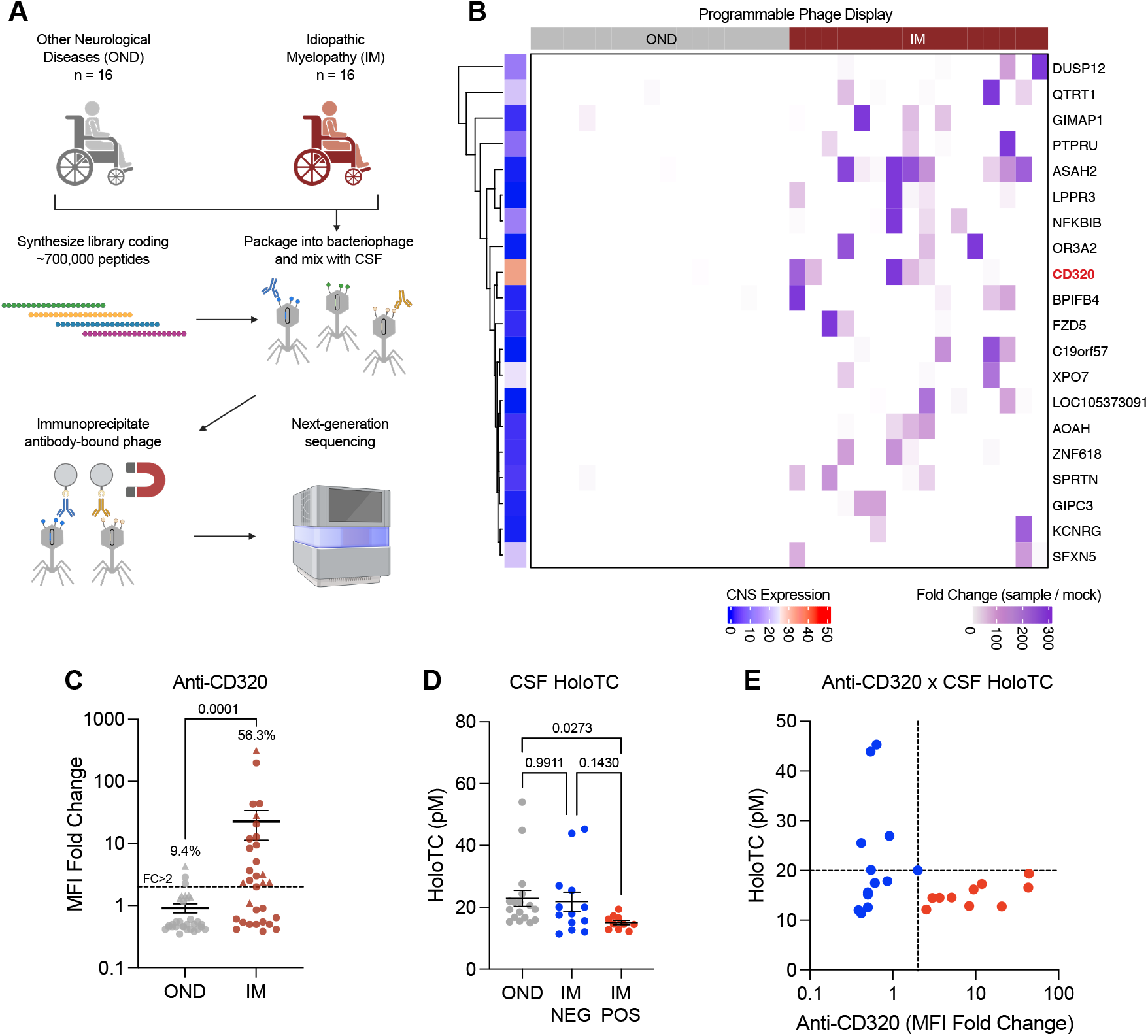
Proteome-wide autoantibody discovery screen identifies anti-CD320 in IM. (**A**) Schematic of PhIP-Seq experiment. (**B**) Heatmap of significantly enriched autoantibody targets in IM cases versus OND controls. The color-coded tile on the left represents the expression of each protein in the CNS (reference: Human Protein Atlas). (**C**) Targeted Luminex-based immunoassay for anti-CD320 in OND controls and IM cases. Anti-CD320 was detected in 3 out of 32 (9.4%) OND controls and 18 out of 32 (56.3%) IM cases (P = 0.0001, two-sided Fisher’s exact test). Circles represent CSF samples and triangles represent serum samples, which are balanced between OND and IM groups. (**D**) CSF bioactive B12 (holoTC) concentration in OND controls, anti-CD320 negative IM cases, and anti-CD320 positive IM cases measured by ELISA (*P* = 0.0273, Welch’s analysis of variance (ANOVA) with Dunnett’s multiple comparisons test). (**E**) Scatter plot demonstrating anti-CD320 Luminex signal versus CSF holoTC concentration in positive (red) and negative (blue) IM cases.

To validate this finding, we developed an orthogonal targeted immunoassay to detect epitope-specific anti-CD320 (Supplementary Figure 2B) comparable to but more quantitative than a traditional cell-based assay (Supplementary Figure 2C). Following the initial PhIP-Seq screen, 16 additional patients were enrolled using the same inclusion and exclusion criteria, comprising 32 total IM cases in an extended discovery cohort. Additional OND controls were randomly selected to balance the groups (MS, n = 11; neurosarcoidosis, n = 1; Leber’s hereditary optic neuropathy, n = 1; migraine, n = 1; B12 deficiency, n = 1; healthy, n = 1; Supplementary Table 2). Anti-CD320 was present in 18 out of 32 IM cases (56.3%) compared to 3 out of 32 OND controls (9.4%, *P* = 0.0001, Figure 2C).

To assess potential metabolic consequences of this autoantibody, we measured the bioactive form of B12 bound to its carrier protein (holotranscobalamin, holoTC) in the CSF of OND controls and IM cases.^16^ HoloTC concentration was decreased in the CSF of anti-CD320 positive IM cases compared to OND controls (*P* = 0.0273, Figure 2D). All anti-CD320 positive cases and several anti-CD320 negative cases demonstrated a low CSF holoTC concentration compared to previously reported normal values (Figure 2E).^17,18^ MMA, a metabolite that accumulates in B12 deficiency, was negatively correlated with holoTC in the CSF of anti-CD320 positive IM cases (*P* = 0.0432), but not in anti-CD320 negative IM cases or OND controls (Supplementary Figure 2D).

There were no statistically significant differences in basic demographic characteristics between anti-CD320 negative and positive IM cases (Table 1, Supplementary Table 3). However, anti-CD320 positive IM cases demonstrated a higher frequency of subacute time course (56% vs 7%, *P* = 0.008) and normal CSF profile (83% vs 50%, *P* = 0.044, Table 1). Although most IM cases demonstrated dorsal or lateral cord signal abnormalities irrespective of antibody status (Figure 3A), combined dorsal and lateral cord abnormalities were more common in anti-CD320 positive cases (61% vs 7%, *P* = 0.003, Table 1). This pattern resembled subacute combined degeneration (SCD), a paradigmatic finding in classic B12 deficiency characterized by demyelination in the dorsal columns and lateral corticospinal tracts of the spinal cord (Figure 3B; Supplementary Figures 3A-F).^19,20,21^ Antecedent viral infections were common but similarly distributed between anti-CD320 negative (21%) and positive (28%) IM cases. Comorbid autoimmune conditions and HIV were also common but similarly distributed (Table 1).

**Figure 3.**
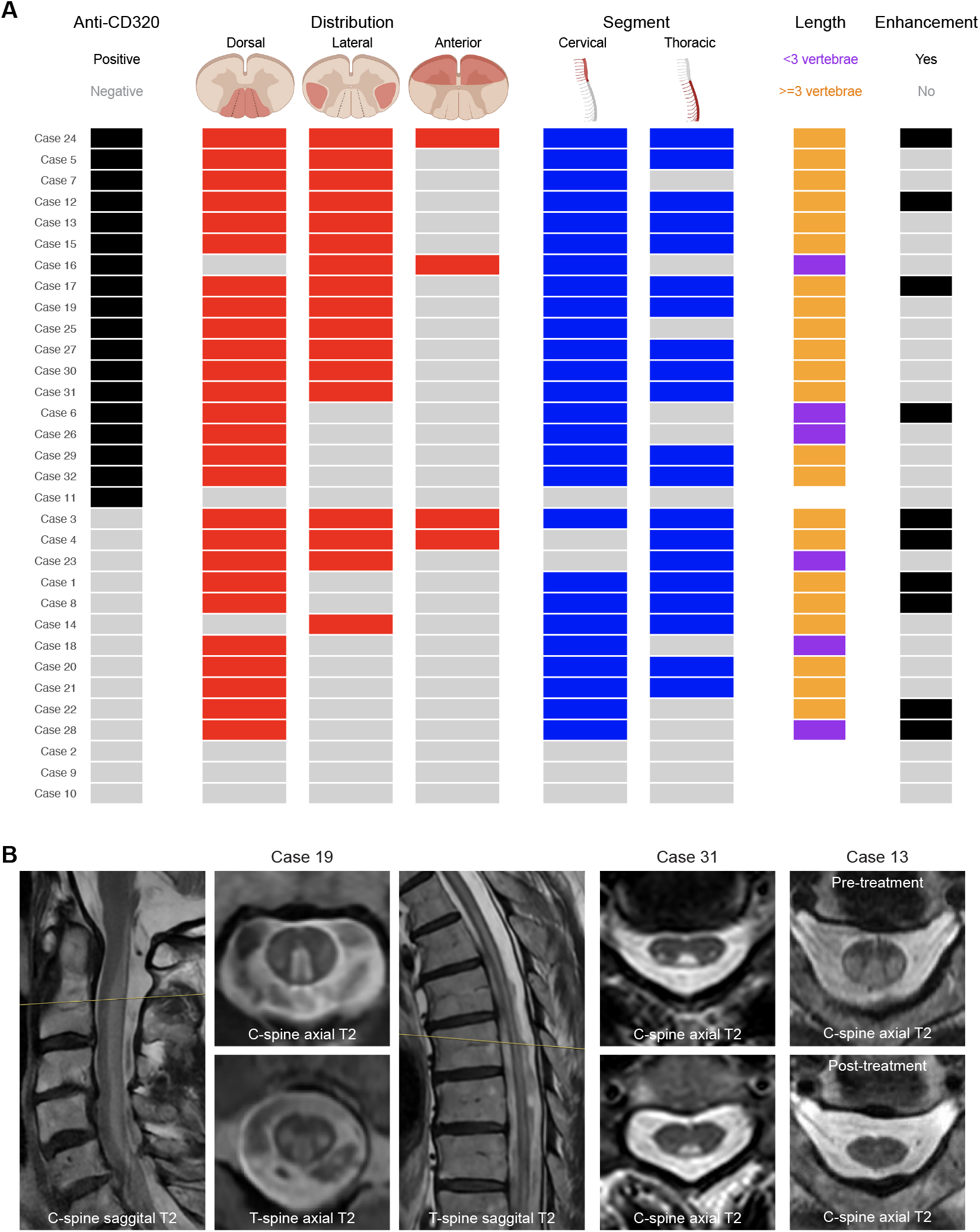
MRI characteristics in anti-CD320 negative and positive IM cases. (**A**) T2 hyperintensity cross-sectional distribution (red), segment localization (blue), length (orange/purple), and presence of contrast-enhancement (black/grey) in anti-CD320 positive (top) and anti-CD320 negative (bottom) IM cases.^22^ (**B**) Representative MRI findings in IM cases with dorsolateral T2 hyperintensities resembling SCD. Pre-treatment (top) and post-treatment (bottom) MRIs in Case 13 demonstrate resolution of cord signal abnormalities after B12 supplementation and rituximab treatment.

To determine if the high prevalence of anti-CD320 in IM is a generalizable finding, we retrospectively collected an independent validation cohort (validation cohort 1) comprising 94 cases of IM using the same selection criteria as the discovery cohort (Supplementary Figure 4A). Validation cohort 1 was slightly younger (mean age 46 versus 54) and contained a higher percentage of female subjects (55% versus 41%) compared to the discovery cohort (Supplementary Tables 4 and 5). We detected anti-CD320 in 43 out of 94 IM cases (45.7%, Figure 4A). CSF holoTC was decreased in anti-CD320 positive cases compared to anti-CD320 negative cases (*P* = 0.0426, Figure 4B) and negatively correlated with anti-CD320 concentration (*P* = 0.0204, Figure 4C).

**Figure 4.**
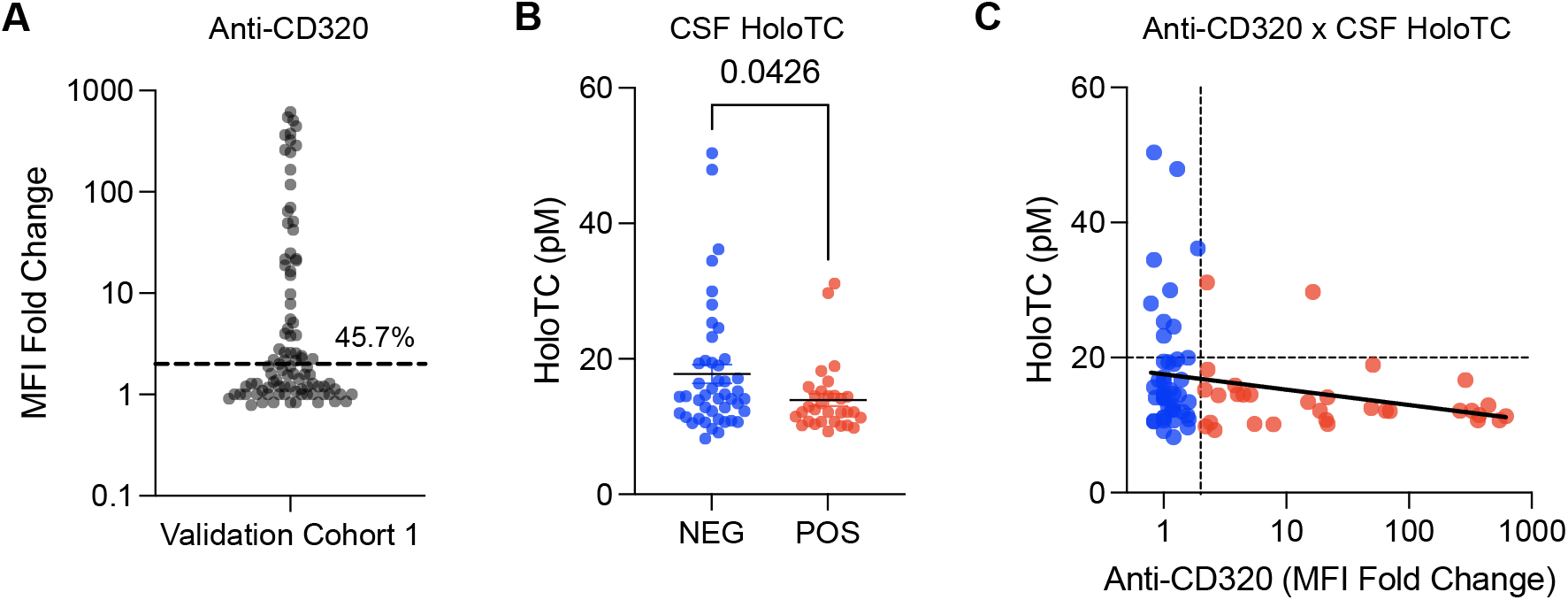
Anti-CD320 status and CSF holoTC concentration in validation cohort 1. (**A**) Targeted Luminex-based immunoassay detected anti-CD320 in 43 out of 94 (45.7%) IM cases. (**B**) CSF holoTC concentration in anti-CD320 negative (blue) and positive (red) IM cases measured by ELISA (P = 0.0426, two-sided Student’s t-test). (**C**) Scatter plot demonstrating anti-CD320 Luminex signal versus CSF holoTC concentration in anti-CD320 negative (blue) and positive (red) IM cases from validation cohort 1. Linear regression line shown in black (*P* = 0.0204, Spearman correlation).

Paired serum and CSF samples were available in a subset of validation cohort 1 (n = 38). In these cases, serum anti-CD320 correlated with CSF anti-CD320 (*P* < 0.0001, Supplementary Figure 4B) but was present at 40-500x higher concentrations (Supplementary Figure 4C). We measured the ratio of CSF holoTC to serum holoTC as a proxy for B12 transport across the blood-brain barrier. Both serum anti-CD320 and CSF anti-CD320 were negatively correlated with this ratio, indicative of impaired B12 transport from blood to CNS (Supplementary Figure 4D). Next, we assessed anti-CD320 dynamics in 9 cases with longitudinal samples available. Anti-CD320 levels remained stable over time (1 day - 7 years follow up, Supplementary Figure 4E), without any cases of seroconversion (negative to positive) or seroreversion (positive to negative).

Comorbid autoantibodies are present in up to 50% of patients with anti-AQP4+ NMOSD,^23^ and antecedent viral-like prodromes are present in up to 61% of patients with anti-MOG+ myelitis.^24^ Given the high frequency of autoimmunity and antecedent viral infection in our discovery cohort of IM, we investigated whether comorbid anti-CD320 is detectable in these other known etiologies of autoimmune myelopathy. To do so, we retrospectively assembled a second independent validation cohort (validation cohort 2) comprising 23 cases of anti-AQP4+ myelitis, 7 cases of anti-MOG+ myelitis, and 25 cases of double anti-AQP4/anti-MOG negative IM. Anti-CD320 was detectable in 17.4% of anti-AQP4+ myelitis, 28.6% of anti-MOG+ myelitis, and 48.0% of IM (Supplementary Figure 5A, Supplementary Tables 4 and 6). CSF was not available for holoTC measurement from anti-AQP4+ or anti-MOG+ myelitis cases.

Seventeen IM patients in the discovery cohort received empiric treatment. Five anti-CD320 negative IM patients were treated with one or multiple immunosuppressive medications, and one out of five clinically improved (Supplementary Table 3). Ten anti-CD320 positive IM patients received immunosuppression, and seven out of ten clinically improved. We previously reported Case 13, man in his 80s presenting after COVID-19 infection with SCD who improved with rituximab and B12 supplementation despite a normal serum B12 level.^7^ In addition to Case 13, four other anti-CD320 positive IM cases received systemic B12 supplementation. Case 27 (subacute worsening on chronic time course) was unresponsive to intravenous immunoglobulin, steroids, plasma exchange, cyclophosphamide, mycophenolate mofetil, and intramuscular B12 supplementation (Supplementary Table 3). Case 19 (subacute time course) transiently improved with steroids and intramuscular B12 injections but later regressed (Supplementary Appendix, Case Vignettes). Case 25 (subacute time course) and Case 26 (subacute time course) improved on B12 supplementation alone (Supplementary Appendix, Case Vignettes).

## Discussion

We identified a known functional autoantibody (anti-CD320) in a substantial proportion of IM cases. Although classic serum B12 deficiency had been ruled out in these patients, CSF bioactive B12 (holoTC) concentration was reduced, consistent with ABCD. Anti-CD320 positive IM cases demonstrated a high frequency of subacute time course, dorsolateral MRI abnormalities, and normal CSF profile. In two independent validation cohorts, a similar proportion of IM cases tested positive for anti-CD320. Comorbid anti-CD320 was detectable in a subset of patients with anti-AQP4+ or anti-MOG+ myelitis, albeit at a lower frequency than in IM. Five patients received B12 supplementation with or without immunomodulatory treatment, and four out of five clinically improved. These findings uncover a potential new etiological contributor to IM that may be amenable to therapeutic intervention.

Classic systemic B12 deficiency is a heterogeneous condition that can remain subclinical or affect any level of the neuroaxis, and ABCD mirrors this heterogeneity. In addition to myelopathy, we previously detected anti-CD320 in patients with cognitive impairment, cerebellar ataxia, and neuropsychiatric SLE, and others have reported anti-CD320 in cutaneous vasculitis and scleroderma.^25,26^ Furthermore, approximately 6% of healthy controls harbor anti-CD320, comparable to the seroprevalence of anti-GAD65.^27,28^ Thus, anti-CD320 is not a specific biomarker for myelopathy. However, the marked enrichment of anti-CD320 in IM, the metabolic evidence of CNS-specific B12 deficiency, and the predilection for dorsolateral tract-specific MRI abnormalities suggest that anti-CD320 is an etiological contributor in IM pathogenesis. Therefore, in the correct clinical context (patients with IM, especially those mimicking SCD), screening for anti-CD320 followed by metabolic confirmation of B12 deficiency in the CSF may be warranted but should not curtail an exhaustive diagnostic workup.

In addition to IM, we detected anti-CD320 in a smaller proportion of patients with known autoimmune etiologies of myelopathy (i.e., anti-AQP4 and anti-MOG). While anti-CD320 may be a bystander in NMOSD and MOGAD, it is also possible that anti-CD320 modulates clinical trajectories in these heterogeneous conditions. Indeed, cases of NMOSD overlapping with classic systemic B12 deficiency have been reported.^29,30^ Future studies with larger cohorts and paired CSF will be necessary to determine if ABCD independently contributes to NMOSD and MOGAD disease progression.

Combining research-based autoantibody discovery with CSF metabolite measurement allowed us to detect anti-CD320 positive IM cases with normal or elevated serum B12 concentrations but low CSF holoTC concentrations, indicative of ABCD. However, we also measured a low CSF holoTC concentration in several anti-CD320 negative IM cases. These findings suggest that our epitope-specific serological assay may miss some cases of ABCD (false negative) or that alternative non-anti-CD320 mediated mechanisms of impaired B12 penetration into the CNS exist. Indeed, genetic polymorphisms in CD320 and transcobalamin that modulate B12 transport have been previously described.^31,32,33,34^ Importantly, low CSF holoTC was correlated with elevated MMA only in anti-CD320 positive IM cases, suggesting that the antibody impairs not just B12 transport into the CNS but also its cellular utilization. Anti-CD320 is hypothesized to target both the luminal side of the blood-brain barrier (impairing B12 transport) and neurons and glia (impairing B12 uptake) via receptor internalization. Thus, both serum and CSF autoantibodies may reduce B12 bioavailability in the CNS.

The triad of subacute time course, dorsolateral spinal cord abnormalities, and normal CSF profile in anti-CD320 positive IM cases mimics the clinical presentation of SCD caused by classic systemic B12 deficiency. Several other conditions can also mimic SCD, including copper deficiency, folate deficiency, vitamin E deficiency, nitrous oxide toxicity, syphilis, and human T-lymphotropic virus 1 (HTLV1) infection.^35^ While these etiologies were ruled out when clinically indicated in the discovery cohort, consideration of all SCD mimics remains an important and challenging component in the workup of IM. Comorbid HIV infection was present in several patients but similarly distributed between anti-CD320 negative and positive cases. Because vacuolar degeneration in HIV myelopathy is pathologically indistinguishable from that of B12 deficiency,^36^ the driving etiology in anti-CD320 and HIV double-positive IM cases remains uncertain. It is possible that HIV and B12 deficiency disturb the same pathways in white matter, but future mechanistic studies will be necessary to explore this clinicopathological phenomenon.

This study has several limitations. First, despite the comparable frequency of anti-CD320 in discovery (n = 32) and validation (n = 94; n = 25) cohorts, this study may not accurately represent the prevalence and clinical characteristics of ABCD in IM at a population level. Second, our focus on anti-CD320 does not preclude the possibility that alternative occult co-morbid etiologies contribute to myelopathy in these patients. For example, our PhIP-Seq experiment revealed several other uncharacterized autoantibodies enriched in IM that warrant further investigation in future studies. Furthermore, emerging rare causes of tract-specific myelopathies such as pegivirus-associated encephalomyelitis or *Mycobacterium haemophilum*-related myelitis were not excluded.^37,38^ Seroreversion of certain known myelitis-associated antibodies can occur.^39,40,41^ Whether these situations account for IM cases in our cohort is not known. Finally, the treatment response to B12 supplementation in anti-CD320 positive IM patients remains anecdotal and potentially confounded by concurrent immunosuppressive treatments; future prospective controlled clinical studies will be necessary to determine therapeutic efficacy. In conclusion, ABCD (anti-CD320 + metabolic evidence of CNS-restricted B12 deficiency) is associated with a substantial proportion of IM, especially in cases with features of SCD despite a normal serum B12 level. Screening for ABCD, either as a primary etiology or a comorbid condition, may improve the diagnosis and treatment of patients with myelopathy.

## Data Availability

All data produced in the present work are contained in the manuscript

## Acknowledgements

We thank Chloe Gerungan for laboratory management, Margherita Vacca for coordination, members of the Wilson lab for discussion, and the research subjects and their families for participation in this study. This work was supported by NINDS (U01NS120836 to M.R.W., S.J.P., J.M.G., A.J.G.; R35NS111644 to S.L.H and M.R.W.; RM1NS138808 to S.J.P., M.R.W., J.L.D.; UE5NS070680 to J.V.P.; K08NS142575 to J.V.P.), the Westridge Foundation (to M.R.W. and A.J.G.), the Burroughs Wellcome Fund (to J.V.P.), and Arc Institute (to J.V.P.). The ORIGINS study is supported by NIH/NINDS (R35NS111644), NMSS (SI-2001-35701), and the Valhalla Foundation.

