## Supplementary Appendix for "Anti-CD320 Autoantibodies and Central Nervous System Vitamin B12 Deficiency in Idiopathic Myelopathy"

**Supplementary Methods**

*Proteome-wide phage display*

We adapted a previously published protocol for phage immunoprecipitation sequencing (PhIP-Seq).[1,2,3] A library containing 731,724 49–amino acid peptides with 25–amino acid overlaps was cloned into T7 bacteriophage. Patient CSF was incubated with $10^{10}$ plaque-forming units of the phage library, antibodies were immunoprecipitated with protein A/G magnetic beads, and antibody-bound phage was amplified in *Escherichia coli* before a second round of immunoprecipitation. Enriched phage lysates were adaptor-ligated and barcoded before pair-end sequencing on an Illumina NovaSeq to a depth of 2 million reads per sample. Reads were trimmed, aligned at the amino acid level using RAPSearch, and normalized to sequencing depth to generate reads per 100,000 (RPK) for each sample. Enriched peptides were identified by calculating the fold change (FC) of normalized counts between samples immunoprecipitated with CSF versus magnetic beads only. Z-scores were calculated for each IM case using all OND controls as a reference and for each OND control using all other OND controls as a reference. Autoantibody "hits" were selected using criteria adapted from a previously published methodology[4]: 1) Z-score > 2 in at least 10% of cases, 2) Z-score > 2 in 0 controls, and 3) FC > 50 in at least 2 cases. A step-by-step protocol is available at www.protocols.io/view/scaled-moderate-throughput-multichannel-phip-proto-8epv5zp6dv1b/v1.

**Supplementary Figures**

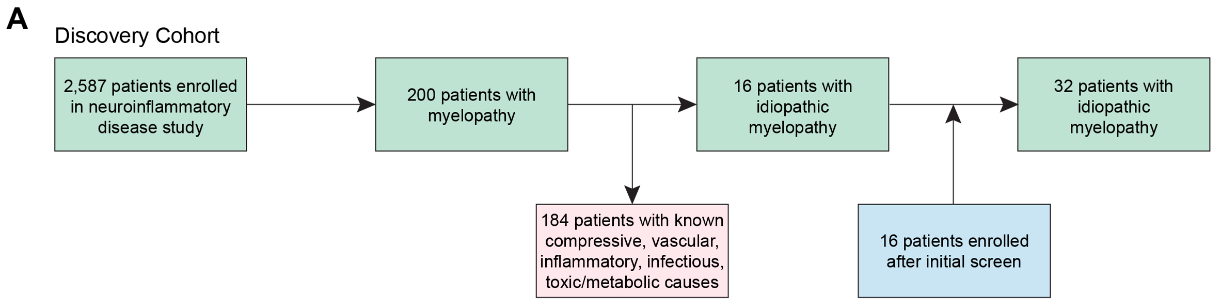

**Supplementary Figure 1.** (**A**) Flow chart summarizing the selection of the discovery cohort. The initial discovery cohort comprised 16 patients with IM. The extended discovery cohort added 16 more patients with IM using the same selection criteria.

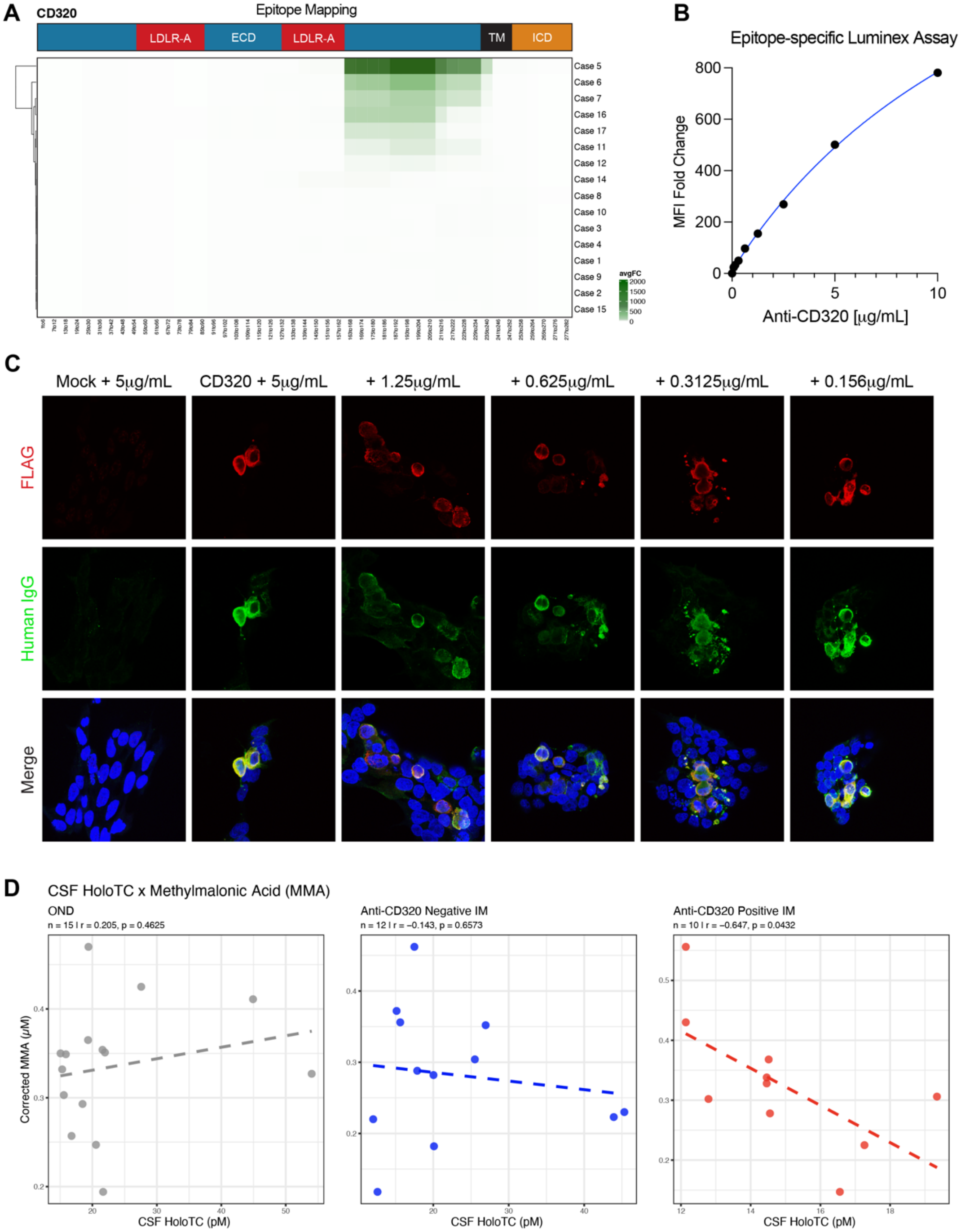

**Supplementary Figure 2**. (**A**) Epitope mapping of anti-CD320 autoantibodies in IM cases. All anti-CD320 positive IM cases enriched the same region in the extracellular domain (ECD, blue) of CD320. (**B**) Standard curve of the epitope-specific Luminex-based immunoassay using a patient-derived monoclonal anti-CD320 antibody. The MFI fold change represents autoantibody enrichment for the antigenic peptide of CD320 conjugated to bovine serum albumin (BSA) versus enrichment for BSA alone. Fitting with an asymmetric sigmoidal 5-parameter logistic curve is shown in blue. (**C**) Representative immunofluorescent images of a cell-based assay using a patient-derived monoclonal anti-CD320 antibody. Compared to mock transfected HEK293T cells (column 1), HEK293T cells transfected with FLAG-tagged CD320 (columns 2-6) demonstrate strong overlap (bottom row) between FLAG signal (red, top row) and human IgG signal (green, middle row) across a range of antibody concentrations. (**D**) Correlation between CSF holoTC concentration and CSF MMA concentration in OND controls (left), chronic/subacute anti-CD320 negative IM cases (middle), or chronic/subacute anti-CD320 positive IM cases (right). Time course is defined as acute (<1 month), subacute (1 month – 6 months), or chronic (>6 months).

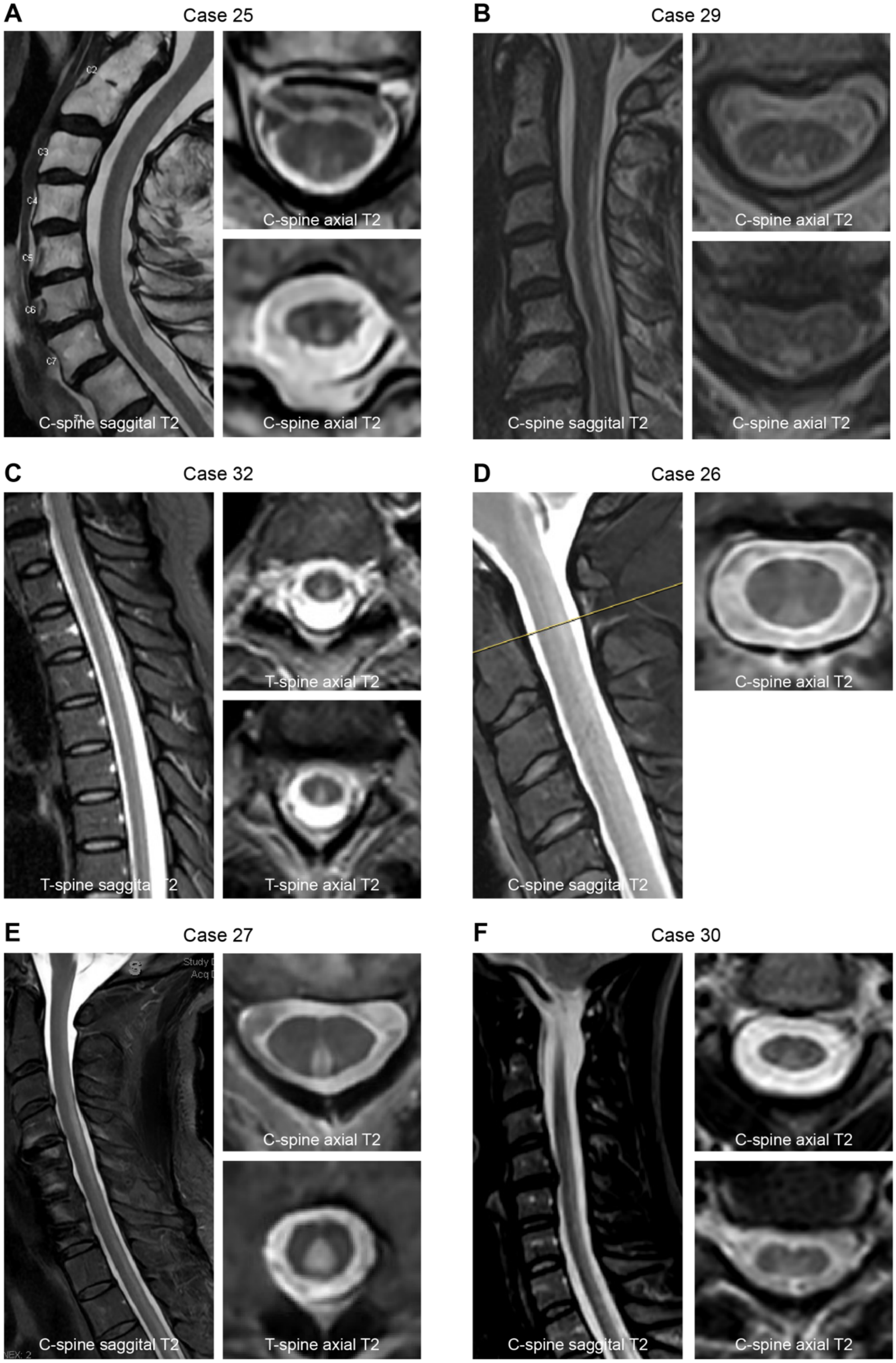

**Supplementary Figure 3.** Representative MRI findings in Case 25 (**A**), Case 29 (**B**), Case 32 (**C**), Case 26 (**D**), Case 27 (**E**), and Case 30 (**F**), demonstrating dorsal and or lateral T2 hyperintensities in the cervical spine and/or thoracic spine.

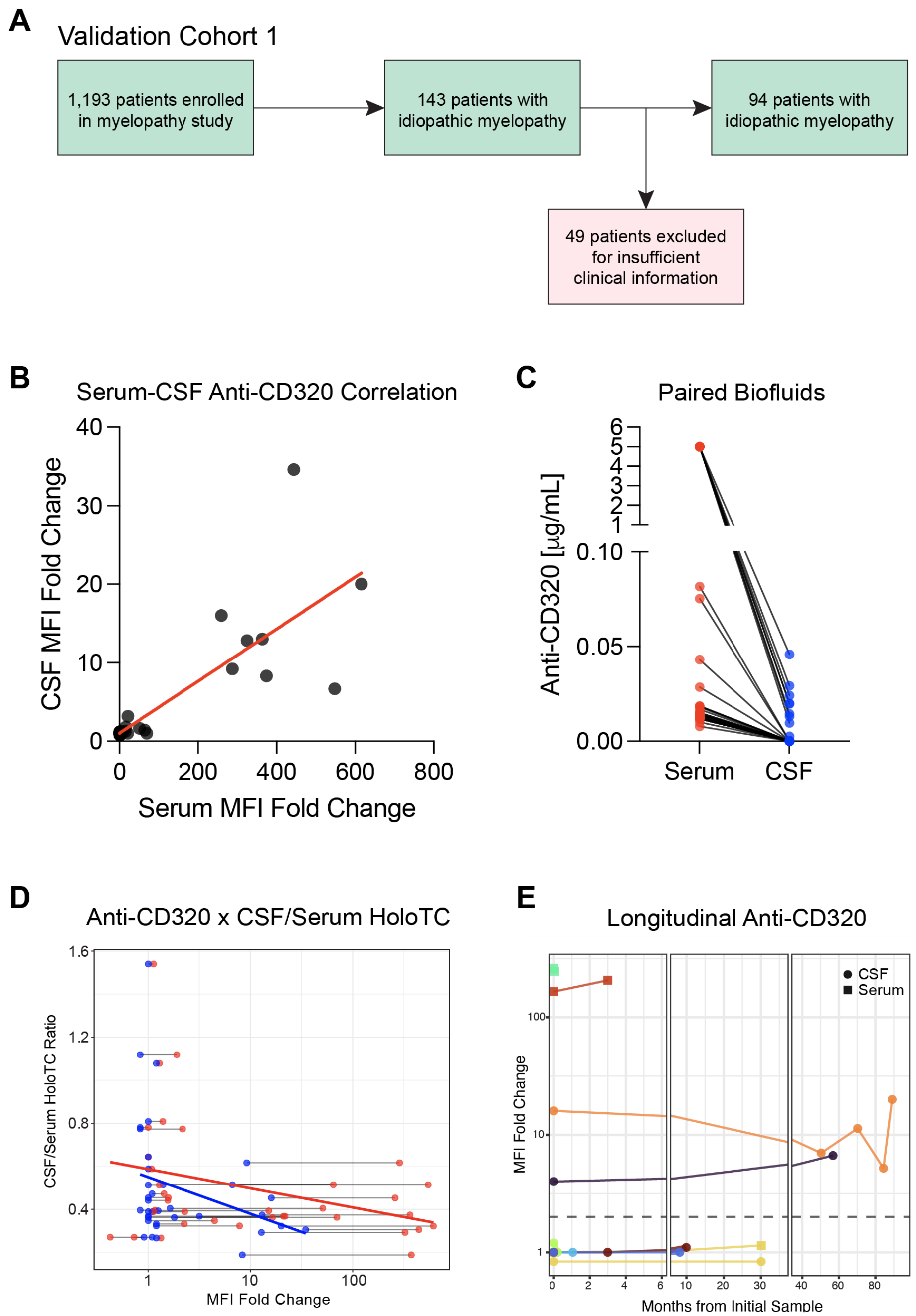

**Supplementary Figure 4**. (**A**) Flow chart summarizing the selection of validation cohort 1. (**B**) Correlation between serum and CSF anti-CD320 MFI fold change in paired samples from validation cohort 1 (n=38, $P < 0.0001$, Pearson correlation). (**C**) Anti-CD320 concentration (interpolated and corrected for dilution factor) in paired serum (red) and CSF (blue) samples. Paired samples are connected by a black line. (**D**) Correlation between anti-CD320 MFI fold change and the CSF/serum holoTC ratio, a proxy for B12 transport from the blood to CNS. Both serum (red, $r = -0.248$, Pearson correlation coefficient) and CSF (blue, $r = -0.24$, Pearson correlation coefficient) anti-CD320 MFI fold changes are negatively correlated with B12 transport. (**E**) Longitudinal anti-CD320 signal in 9 IM cases from Validation Cohort 1.

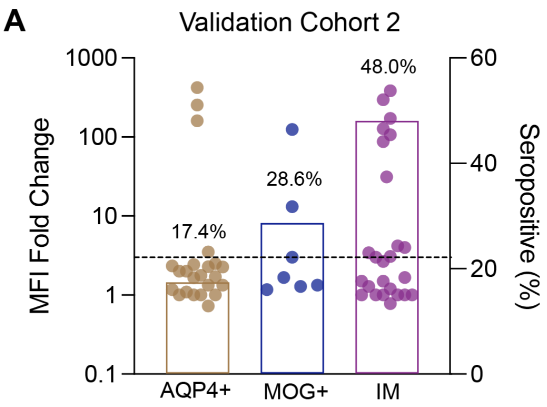

**Supplementary Figure 5**. (**A**) Anti-CD320 MFI fold change (left y-axis) and seropositive rate (right y-axis) in anti-AQP4+ myelopathy (brown), anti-MOG+ myelopathy (blue), and IM (purple) cases from validation cohort 2.

### Supplementary Tables

| Antigen | Subcellular Localization | Tissue Specificity |
|---|---|---|
| AOAH | secreted | Lymphoid tissue - Immune response |
| ASAH2 | intracellular | Small intestine - Absorption |
| BPIFB4 | secreted | Pituitary gland - Hormone signaling |
| C19ORF57 | intracellular | Testis - Spermatogenesis |
| CD320 | membrane | Brain - Mitochondrial translation |
| DUSP12 | intracellular | Non-specific - Transcription |
| FZD5 | membrane | Liver & Intestine - Lipid metabolism |
| GIMAP1 | membrane | Lymphoid tissue - Immune response |
| GIPC3 | intracellular | Retina & Testis - Cilium |
| KCNRG | intracellular | Fallopian tube - Tissue development |
| LPPR3 | membrane | Brain - Neuronal signaling |
| NFKBIB | intracellular | Testis - Spermatogenesis |
| OR3A2 | intracellular\|membrane | Brain - Olfactory receptor |
| PTPRU | membrane | Non-specific - Unknown function |
| QTRT1 | intracellular | Non-specific - Protein processing |
| SFXN5 | intracellular | Brain - Neuronal signaling |
| SPRTN | intracellular | Testis - Basic cellular processes |
| XPO7 | intracellular\|membrane | Non-specific - Transcription |
| ZNF618 | intracellular | Thyroid gland - Unknown function |

**Supplementary Table 1**. Summary of PhIP-seq hits enriched in IM. Most hits demonstrate intracellular localization or non-CNS tissue specificity based on data from the Human Protein Atlas RNA-seq expression data. CD320, LPPR3, and OR3A2 were the only hits with membrane localization and brain specificity. We focused on CD320 because autoantibodies targeting this antigen were previously reported in a case of IM.

| | OND | IM |
|---|---|---|
| **No. of patients** | 32 | 32 |
| **Age (range)** | 35 (19-54) | 54 (26-80) |
| **Sex (%)** | | |
| Female | 21 (66) | 13 (41) |
| Male | 11 (34) | 19 (59) |

**Supplementary Table 2. Comparison of OND controls and IM cases in discovery cohort.**

*Removed per MedRxiv policy regarding detailed description of comorbidities.*

**Supplementary Table 3. Clinical summaries of the discovery cohort.** Anti-CD320 positive cases are highlighted in red. Serum B12 was borderline low (280) in Case 24 but with normal homocysteine and MMA. Serum B12 was normal (404) on initial presentation in Case 27 and borderline low (249) on second presentation then repleted (1378).

| | Discovery Cohort | Validation Cohort 1 | Validation Cohort 2 |
|---|---|---|---|
| **No. of patients** | 32 | 94 | 25 |
| **Age (range)** | 54 (26-80) | 46 (18-78) | 58 (31-76) |
| **Sex (%)** | | | |
| Female | 13 (41) | 52 (55) | 9 (36) |
| Male | 19 (59) | 42 (45) | 16 (64) |
| **Anti-CD320 (%)** | | | |
| Negative | 14 (44) | 51 (54) | 13 (52) |
| Positive | 18 (56) | 43 (46) | 12 (48) |

**Supplementary Table 4. Comparison of discovery and validation cohorts.**

| Validation Cohort 1 | Anti-CD320 negative | Anti-CD320 positive | *P*-value |
|---|---|---|---|
| **No. of patients (%)** | 51 (54) | 43 (46) | |
| **Age (range)** | 46 (18-78) | 45 (20-71) | 0.624 |
| **Sex (%)** | | | 0.680 |
| Female | 27 (53) | 25 (58) | |
| Male | 24 (47) | 18 (42) | |
| **CSF findings (%)** | | | 0.347 |
| Normal | 21 (48) | 19 (61) | |
| Inflammatory | 23 (52) | 12 (39) | |

**Supplementary Table 5. Comparison of anti-CD320 negative and positive IM cases in validation cohort 1.** Age was compared using a two-sided Student's t-test. Sex and CSF findings were compared using a two-sided Fisher's exact test.

| Validation Cohort 2 | Anti-CD320 negative | Anti-CD320 positive | *P*-value |
|---|---|---|---|
| **No. of patients (%)** | 13 (52) | 12 (48) | |
| **Age (range)** | 61 (34-76) | 54 (31-76) | 0.188 |
| **Sex (%)** | | | 0.999 |
| Female | 5 (38) | 4 (33) | |
| Male | 8 (62) | 8 (67) | |
| **MRI pattern (%)** | | | 0.431 |
| Normal | 2 (15) | 1 (8) | |
| Dorsolateral | 2 (15) | 5 (42) | |
| Other | 9 (70) | 6 (50) | |

**Supplementary Table 6. Comparison of anti-CD320 negative and positive IM cases in validation cohort 2.** Age was compared using a two-sided Student's t-test. Sex and MRI findings were compared using a two-sided Fisher's exact test.

---

[1] Larman, H. B. *et al.* Autoantigen discovery with a synthetic human peptidome. *Nat. Biotechnol.* **29**, 535–541 (2011).

[2] Mandel-Brehm, C. *et al.* Kelch-like Protein 11 Antibodies in Seminoma-Associated Paraneoplastic Encephalitis. *N. Engl. J. Med.* **381**, 47–54 (2019).

[3] Vazquez, S. E. *et al.* Identification of novel, clinically correlated autoantigens in the monogenic autoimmune syndrome APS1 by proteome-wide PhIP-Seq. *Elife* **9**, (2020).

[4] Vazquez SE, Mann SA, Bodansky A, et al. Autoantibody discovery across monogenic, acquired, and COVID-19-associated autoimmunity with scalable PhIP-seq. Elife [Internet] 2022;11. Available from: http://dx.doi.org/10.7554/eLife.78550
